# Detecting corticospinal tract impairment in tumor patients with fiber density and tensor-based metrics

**DOI:** 10.1101/2020.10.28.20220293

**Authors:** L. S. Fekonja, Z. Wang, D. B. Aydogan, T. Roine, M. Engelhardt, F. R. Dreyer, P. Vajkoczy, T. Picht

**Author notes:** Corresponding author: Lucius S. Fekonja,Address: Charité Universitätsmedizin Berlin, Klinik für Neurochirurgie mit Arbeitsbereich Pädiatrische, Neurochirurgie, Campus Charité Mitte, Luisenstraße 64, 10117 Berlin.

## Abstract

Tumors infiltrating the motor system lead to significant disability, often caused by corticospinal tract injury. The delineation of the healthy-pathological white matter (WM) interface area, for which diffusion magnetic resonance imaging (dMRI) has shown promising potential, may improve treatment outcome. However, up to 90% of white matter (WM) voxels include multiple fiber populations, which cannot be correctly described with traditional metrics such as fractional anisotropy (FA) or apparent diffusion coefficient (ADC). Here, we used a novel fixel-based along-tract analysis consisting of constrained spherical deconvolution (CSD) based probabilistic tractography and fixel-based apparent fiber density (FD), capable of identifying fiber orientation specific microstructural metrics. We addressed this novel methodology’s capability to detect corticospinal tract impairment.

We measured and compared tractogram-related FD and traditional microstructural metrics bihemispherically in 65 patients with WHO grade III and IV gliomas infiltrating the motor system. The cortical tractogram seeds were based on motor maps derived by transcranial magnetic stimulation. We extracted 100 equally distributed cross-sections along each streamline of corticospinal tract (CST) for along-tract statistical analysis. Cross-sections were then analyzed to detect differences between healthy and pathological hemispheres.

All metrics showed significant differences between healthy and pathologic hemispheres over the entire tract and between peritumoral segments. Peritumoral values were lower for FA and FD, but higher for ADC within the entire cohort. FD was more specific to tumor-induced changes in CST than ADC or FA, whereas ADC and FA showed higher sensitivity.

The bihemispheric along-tract analysis provides an approach to detect subject-specific structural changes in healthy and pathological WM. In the current clinical dataset, the more complex FD metrics did not outperform FA and ADC in terms of describing corticospinal tract impairment.

## 1. Introduction^1^

In previous studies we introduced the combination of navigated transcranial magnetic stimulation (TMS) cortical motor mapping and tractography to improve surgery of motor eloquent brain tumors (Krieg et al., 2016; Lefaucheur & Picht, 2016; Picht, Frey, Thieme, Kliesch, & Vajkoczy, 2016; Rosenstock, Grittner, et al., 2017). In a recent study we could also demonstrate that the segmental analysis of diffusion tensor imaging (DTI) derived metrics, such as fractional anisotropy (FA) and apparent diffusion coefficient (ADC), correlated with clinical outcomes (Rosenstock, Giampiccolo, et al., 2017). Here, we now set out to investigate whether more complex metrics derived from spherical deconvolution and probabilistic tractography, which allow for more detailed analysis of the white matter, would prove superior in terms of detecting tumor induced white matter (WM) changes. In this context we analyzed the structural impact of gliomas affecting the corticospinal tract (CST) in 65 patients. This was carried out without the generation of a group template because of the lateralized pathology, which allows a clear deduction of interhemispheric differences on the subject-level (D. A. Raffelt et al., 2015). We compared the pathological with the healthy hemisphere and focused on describing tumor-induced changes along the CST with dMRI. We used CSD-based probabilistic tractography at an individual scale within the MRtrix3 framework (Tournier et al., 2019).

DTI enables quantification of the molecular diffusion rate, ADC, or the directional preference of diffusion, FA (Soares, Marques, Alves, & Sousa, 2013). ADC and FA are established metrics integrated as predictive features in neurosurgical studies (Rosenstock, Giampiccolo, et al., 2017). The two main diffusion tensor-derived parameters, ADC and FA, are based on voxel-wise eigenvalues, which represent the magnitude of the diffusion process in the principal diffusion orientation and two directions perpendicular to it. These values are influenced by different factors (Colby et al., 2012). ADC is a measure of the overall diffusivity in a single voxel, regardless of its orientation. It is higher where water diffuses more easily, e.g. in ventricles, lower in structures with high tissue density and consequently more diffusion barriers, such as GM (Van Hecke, Emsell, & Sunaert, 2016). FA describes the directional coherence of water diffusion in tissue and is modulated by numerous biological factors, such as the microstructural and architectural organization of white matter, myelination and non-white matter partial volume effects. Further influences on FA modulation are methodological factors, such as the choice of the estimation, preprocessing methods and subjective selection of regions of interests (ROIs) (Roine et al., 2014; Veraart, Sijbers, Sunaert, Leemans, & Jeurissen, 2013).

In contrast to DTI, CSD can distinguish complex fiber populations in the brain. In brief, CSD estimates fiber orientation distributions (FODs) within each voxel, based on the expected signal from a single collinearly oriented fiber population (Tournier, Calamante, Gadian, & Connelly, 2004). By leveraging the rich information in FODs, probabilistic tractography algorithms, such as the iFOD2, have been proposed to address limitations of tensor-based tractography methods (Tournier, Mori, & Leemans, 2011). In up to 90% of all WM voxels, multiple fiber orientations were observed, and 30% to 40% of these WM voxels contain more than three fiber populations (Jeurissen, Leemans, Tournier, Jones, & Sijbers, 2013; Riffert, Schreiber, Anwander, & Knösche, 2014; Tournier, 2019; Vos, Jones, Jeurissen, Viergever, & Leemans, 2012). Moreover, non-white matter contamination is present in more than a third of the WM voxels (Roine et al., 2014) and has been addressed by multi-tissue CSD methods (Dhollander, Raffelt, & Connelly, 2016; Jeurissen, Tournier, Dhollander, Connelly, & Sijbers, 2014b; Roine et al., 2015).

A complete picture about the underlying white matter architecture is highly relevant with regard to adequate risk estimation and neurosurgical planning (Mormina et al., 2015). To that end, in addition to the conventional DTI measures, modern CSD-based fiber density (FD) and fixel-based analysis (FBA) methods offer promising opportunities, since they are related to the intra-axonal restricted compartment that is specific to a certain fiber orientation within a voxel (D. A. Raffelt et al., 2017). Based on its advantages for the analysis of crossing fiber regions, we expect this metric to improve the detection of tumor-induced changes along the CST, and obtain more specific information about the microstructural effects of tumors in combination with traditional FA or ADC measures. Furthermore, we expect higher specificity of FD in detecting the peritumoral segments, most importantly at the tumor-white matter interface, which is surgically the most important area. However, the translation of advanced neuroimaging to clinical settings is slow both in terms of adapting modern methods and imaging protocols. While there exist tools to use the modern CSD and probabilistic tractography with conventional images, for tumor patients, little is known about how applicable they prove with existing conventional neuroimaging protocols. Nevertheless, clinical feasibility, robustness, and methodological superiority has been proven (Farquharson et al., 2013; Petersen et al., 2017). Until now, fixel-based studies have concentrated on group analyses without subject-specific examination of tumor patients for neurosurgical planning (D. A. Raffelt et al., 2017). We developed a new variant of FD for the fiber orientation specific along-tract investigation of microstructural properties in relation to infiltrating tumors.

Importantly, we used state-of-the-art TMS methods for motor mapping to find functionally critical regions of interest (ROIs) and used these as seed points to generate streamlines. This approach is shown to be highly effective for surgical planning (Picht et al., 2016), therefore it is superior to studying the whole CST, which lacks information about patient and tumor specific functional consequences of neurosurgery.

## 2. Material & Methods

### 2.1. Ethical standard

The study proposal is in accordance with ethical standards of the Declaration of Helsinki and was approved by the Ethics Commission of the Charité University Hospital (#EA1/016/19). All patients provided written informed consent for medical evaluations and treatments within the scope of the study.

### 2.2 Patient selection

We included n=65 left- and right-handed adult patients in this study (25 females, 40 males, age 55.6±15.2, age range 24-81). Only patients with an initial diagnosis of unilateral WHO grade III & IV gliomas (14 WHO grade III, 51 WHO grade IV) were included (Table 1). All tumors were infiltrating M1 and the CST or implied critical adjacency, either in the left or right hemisphere. Patients with recurrent 143 tumors, previous radiochemotherapy, 144 multicentric or non-glial tumors were not considered. 145

**Table 1.** Patient demographics.

|  | Number (%) |
| --- | --- |
| Demographics |  |
| Sample size | 65 |
| Age | 55.6±15.2 |
| Female | 25(38) |
| Male | 40(62) |
| Glioma Degree |  |
| Glioma III | 14 (22) |
| Glioma IV | 51 (78) |
| Tumor Location |  |
| Frontal | 33 (51) |
| Temporal | 7 (11) |
| Insular | 9 (14) |
| Parietal | 16 (25) |

### 2.3 Image acquisition

MRI data were acquired on a Siemens Skyra 3T scanner (Erlangen, Germany) equipped with a 32-channel receiver head coil at Charité University Hospital, Berlin, Department of Neuroradiology. These data consisted of a high-resolution T1-weighted structural (TR/TE/TI 2300/2.32/900 m s, 9° flip angle, 256 × 256 matrix, 1 mm isotropic voxels, 192 slices, acquisition time: 5 min) and a single shell dMRI acquisition (TR/TE 7500/95m s, 2 × 2 × 2 mm^3^ voxels, 128 × 128 matrix, 60 slices, 3 b 0 volumes), acquired at b = 1000 s/mm^2^ with 40 gradient orientations, for a total acquisition time of 12 minutes.

### 2.4 Preprocessing and processing of MRI data

All T1 images were registered to the dMRI data sets using Advanced Normalization Tools (ANTs) with the Symmetric Normalization (SyN) transformation model (Avants et al., 2011; Grabner et al., 2006). The preprocessing of dMRI data included the following and was performed within MRtrix3 (Tournier et al., 2019) in order: denoising (Veraart et al., 2016), removal of Gibbs ringing artefacts (Kellner, Dhital, Kiselev, & Reisert, 2016), correction of subject motion (Leemans & Jones, 2009), eddy-currents (J. L. R. Andersson et al., 2017) and susceptibility-induced distortions (J. L. Andersson, Skare, & Ashburner, 2003) in FMRIB Software Library (Jenkinson, Beckmann, Behrens, Woolrich, & Smith, 2012), and subsequent bias field correction with ANTs N4 (Tustison et al., 2010). Each dMRI data set and processing step was visually inspected for outliers and artifacts. Scans with excessive motion were initially excluded (over 10% outlier slices). We upsampled the dMRI data to a 1.3 mm isotropic voxel size before computing FODs to increase anatomical contrast and improve downstream tractography results and statistics. To obtain ADC and FA scalar maps, we first used diffusion tensor estimation using iteratively reweighted linear least squares estimator, resulting in scalar maps of tensor-derived parameters (Basser, Mattiello, & LeBihan, 1994; Veraart et al., 2013). For voxel-wise modelling we used a robust and fully automated and unsupervised method. This method allowed to obtain 3-tissue response functions representing single-fiber combined white and grey matter and CSF from our data with subsequent use of multi-tissue CSD to obtain tissue specific orientation distribution functions and white matter FODs (Dhollander et al., 2016; Jeurissen et al., 2014b; Tournier, Calamante, & Connelly, 2007).

### 2.5 Transcranial magnetic stimulation

Non-invasive functional motor mapping of both pathologic and healthy hemispheres was performed in each patient using navigated transcranial magnetic stimulation (nTMS) with Nexstim eXimia Navigated Brain Stimulation (NBS). Briefly, each patient’s head was registered to the structural MRI through the use of anatomical landmarks and surface registration. The composite muscle action potentials were captured by the integrated electromyography unit (EMG) (sampling rate 3 kHz, resolution 0.3 mV; Neuroline 720, Ambu). The muscle activity (motor evoked potential, MEP amplitude ≥ 50 μV) was recorded by surface electrodes on the abductor pollicis brevis and first dorsal interosseous (FDI). First, the FDI hotspot, defined as the stimulation area that evoked the strongest MEP, was determined. Subsequently, the resting motor threshold (RMT), defined as the lowest stimulation intensity that repeatedly elicits MEPs, was defined using a threshold-hunting algorithm within the Nexstim eximia software. Mapping was performed at 105% RMT and 0.25 Hz. All MEP amplitudes > 50 μV (peak to peak) were considered as motor positive responses and exported in the definitive mapping (Picht et al., 2011). The subject-specific positive responses of the FDI were exported as binary 3 × 3 × 3 mm^3^ voxel masks per response in the T1 image space.

### 2.6 Tractography

Probabilistic tractography was performed in each hemisphere with the iFOD2 algorithm by using above mentioned nTMS derived cortical seeding ROI. A second inclusion ROI was defined in the medulla oblongata. Tracking parameters were set to default with a FOD amplitude cutoff value of 0.1, a streamline minimum length of 5 x voxel size and a maximum streamline length of 100 x voxel size. For each tractogram describing the CST, we computed 5000 streamlines per hemisphere. Each streamline of the tractograms was resampled along its length to 100 points. Peritumoral segments were defined in relation to the resampled points within the range 1-100 in all individual tractograms by visual inspection performed by one neuroscientist and one expert neurosurgeon with 4 and 20 years of experience, in that order. Subsequently, values of associated FA, ADC and FD scalar maps were sampled along the derived 100 segments of each streamline (Fig. 1 & 2). The code used for the tractography pipeline is archived as a shell script on Zenodo (https://zenodo.org/record/3732348) and openly accessible (Fekonja et al., 2020).

**Fig. 1.**
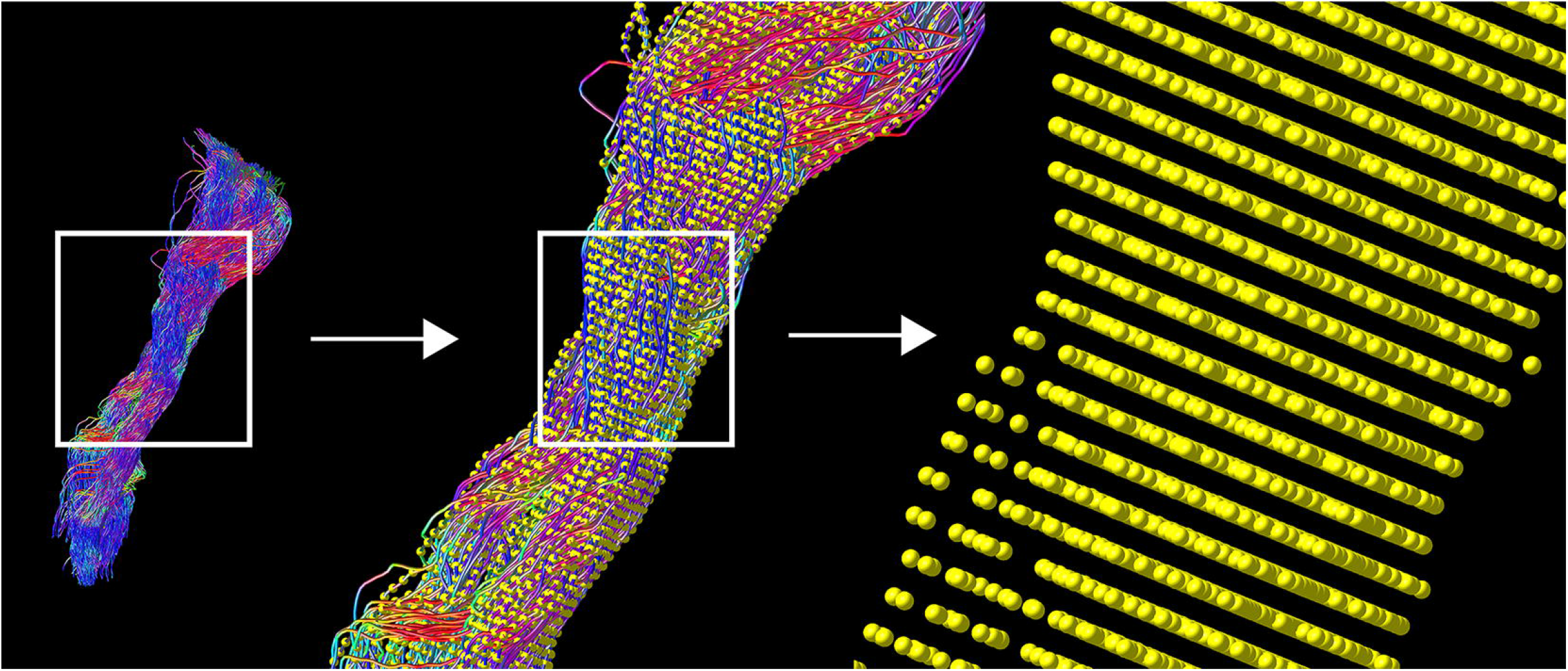
TMS-based tractography of the CST and subsequent along-tract resampling of streamlines. The tractogram shows streamlines in relation to cortical hand representation derived by TMS-ROIs (left). The first zoom shows a combination with resampled points (yellow), overlaid on each streamlines (middle). The second, larger magnification reveals the single points, derived by resampling along the streamlines (right).

**Fig. 2.**
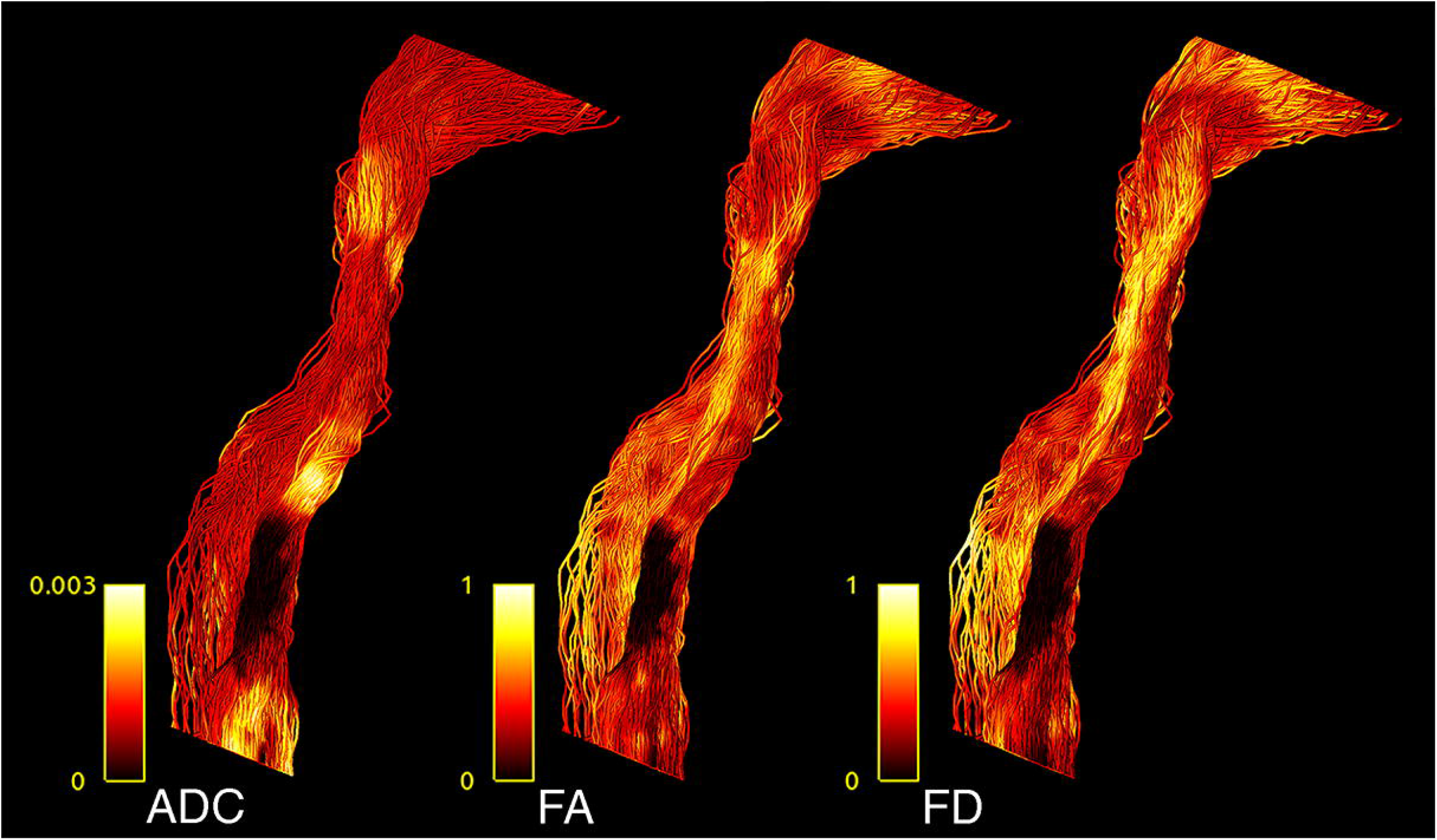
CST tractogram with mapped ADC (left), FA (middle) and FD (right) scalar values, illustrating the methodological differences of scalar maps sampling.

### 2.7 Computation of along-tract FD values using FBA

A fixel is considered as a specific fiber population within a voxel (D. A. Raffelt et al., 2015; D. A. Raffelt et al., 2017). For each subject, segmentations of continuous FODs via the integrals of the FOD lobes were performed to produce discrete fixel maps which are developed to indicate voxel-based measures of axon diameters, weighted by their relative volumes within voxels (D. A. Raffelt et al., 2017; Smith, Tournier, Calamante, & Connelly, 2013). With higher-order diffusion models, such as CSD, parameters related to FD can be extracted for individual fixels (D. A. Raffelt et al., 2015). FBA is able to identify effects in specific fiber pathways and in crossing fibers regions, unlike voxel-based analysis (D. A. Raffelt et al., 2015). After obtaining the fixels for all voxels in an image, FD values along CST tractograms were computed in 4 steps: (i) fixels associated with CSTs were obtained using fixel tract-density imaging, (ii) fixels in the image were thresholded based on the CST fixels which eliminates the contributions of other tracts that are present in these voxels, (iii) the mean FD of the remaining fixels were exported as a scalar image, and (iv) FD values were interpolated along the 100 sampled points of each streamline present in the CST tractograms. The code used for the tract-based fixel image construction pipeline is archived as a shell script on Zenodo (https://zenodo.org/record/3732348) and openly accessible (Fekonja et al., 2020).

### 2.8 Statistical analysis

Confirmatory statistical analysis was performed using RStudio version 1.2.5019 (https://rstudio.com) with R version 3.6.1 (https://cran.r-project.org). We compared FD with traditional tensor-derived ADC and FA to study signal changes between healthy and pathological hemispheres. To analyze the behavior of the different metrics, we used above mentioned resampled streamlines, comparing the median values for each of the 100 CST segments per 5000 streamlines per hemisphere. To model the tumor-related effect on each metric, a linear mixed model (package lmerTest_3.1-0 under R version 3.6.1) was built for each metric using the metric’s value as dependent variable, hemisphere (0, healthy; 1, pathological) as independent variable and a random intercept for subjects (Kuznetsova, Brockhoff, & Christensen, 2017). Thus, each model contained 13000 data points (65 subjects * 2 hemispheres * 100 median tract segment values per streamline). Further, we repeated this analysis for the peritumoral area according to our hypothesis to find stronger effects in these segments. Each of these models contained 4138 data points, with each subject contributing a different number of peritumoral segments depending on tumor location and size. All effects were considered significant using a two-sided p-value of 0.05. All models were examined for patterns in the residuals (deviation from normality via QQ-plots, pattern fitted values vs. residuals). All plots were generated with the ggplot2 library within tidyverse (Wickham, 2009; Wickham et al., 2019). Tests for sensitivity (*n* of true positive predicted segments/*n* of true positive predicted segments + *n* of false negative predicted segments) and specificity (*n* of true negative predicted segments/*n* of true negative predicted segments + *n* of false positive predicted segments) were based on classified tract segments (0 non-tumorous, 1 tumorous) in relation to the obtained significant or non-significant differences between healthy and pathological hemispheres per segment (classified as 0 and 1). These tests were performed with Bonferroni-adjusted alpha levels of 0.0005 (0.05/100) and thresholded only for large effects (≥ 0.474) with Cliff’s delta due to the non-normal distribution. The script used to perform the statistical analysis and produce this manuscript is available on and archived in Zenodo (Fekonja et al., 2020).

### 2.9 Data availability

Parts of the data that support the findings of this study are not publicly available due to information that could compromise the privacy of the research participants but are available from the corresponding author on reasonable request. However, code we have used is openly available under the following address (https://doi.org/10.5281/zenodo.3732348) and is cited at the corresponding passage in the article (Fekonja et al., 2020).

## 3. Results

TMS mapping, the calculation of TMS-ROI-based streamlines and the extraction of ADC, FA and FD were feasible in each subject (cf. Fig. 3) and showed either close tumor-tract distance (< 8mm, n = 3) or adjacency or direct infiltration of the CST by the tumor (n = 62). Visual inspection of boxplots showed differences between pathological and healthy hemispheres for ADC, FA, and FD (Fig 4A). As expected, these differences were larger when looking at the peritumoral area only (Fig 4B). Further, a larger variability in ADC values could be observed in the pathological hemisphere in general and the peritumoral area specifically. When plotting values along the entire CST, distinct patterns of variation between hemispheres could be observed. ADC showed no significant differences in the non-peritumoral segments but showed significant differences in peritumoral segments, even stronger than FA and FD. In contrast, FA and FD values showed differences both in the non-peritumoral and peritumoral segments (Fig. 5, Fig. 6, Table 2). The distribution of tumors along the CST is indicated in Fig. 6. Additionally, the tumor-induced variability in peritumoral ADC values in contrast to the entire CST becomes particularly evident here (Fig. 5). Finally, the information shown in Fig. 2 highlights and visualizes the advantages of FOD representation in regard to multiple fiber populations. The CSD method identifies multiple appropriately oriented fiber populations in a voxel including multiple fiber populations, while the DTI-based method does not represent multiple fiber populations within each voxel and does not provide an orientation estimate corresponding to any of the existing fiber populations (Farquharson et al., 2013), cf. Fig. 3.

**Table 2.**
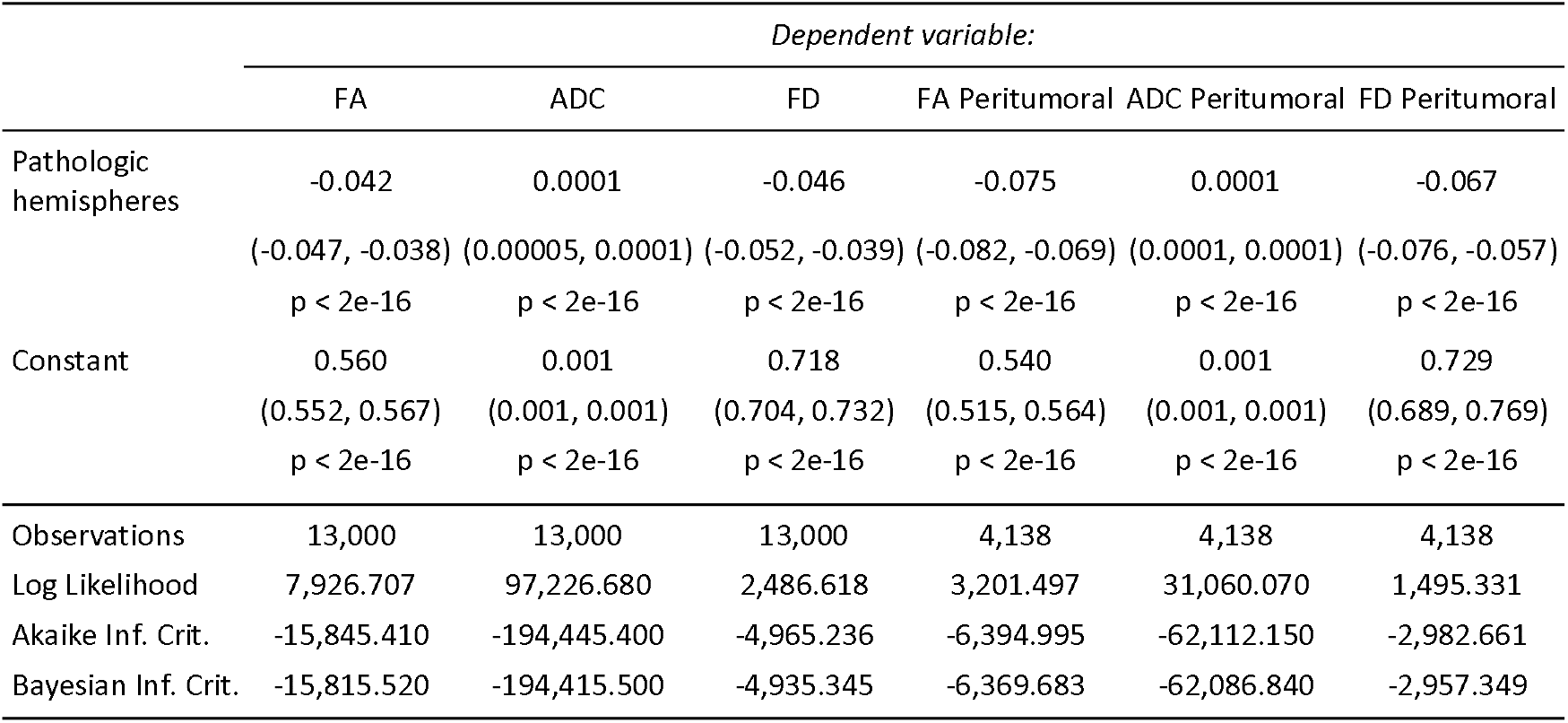
Results of linear mixed model analysis. Models 1-3 show results for the entire CST for FA, ADC and FD, models 4-6 for the peritumoral segments respectively. The table shows regression coefficients for the fixed effect of hemisphere and the intercept with their respective standard error in brackets. Further, number of observations for each model, the log likelihood ratio, Akaike information criterion and Bayesian information criterion are stated.

|  | *Dependent variable:* |  |  |  |  |  |
| --- | --- | --- | --- | --- | --- | --- |
|  | FA | ADC | FD | FA Peritumoral | ADC Peritumoral | FD Peritumoral |
| Pathologic hemispheres | -0.042<br>(-0.047, -0.038)<br>p < 2e-16 | 0.0001<br>(0.00005, 0.0001)<br>p < 2e-16 | -0.046<br>(-0.052, -0.039)<br>p < 2e-16 | -0.075<br>(-0.082, -0.069)<br>p < 2e-16 | 0.0001<br>(0.0001, 0.0001)<br>p < 2e-16 | -0.067<br>(-0.076, -0.057)<br>p < 2e-16 |
| Constant | 0.560<br>(0.552, 0.567)<br>p < 2e-16 | 0.001<br>(0.001, 0.001)<br>p < 2e-16 | 0.718<br>(0.704, 0.732)<br>p < 2e-16 | 0.540<br>(0.515, 0.564)<br>p < 2e-16 | 0.001<br>(0.001, 0.001)<br>p < 2e-16 | 0.729<br>(0.689, 0.769)<br>p < 2e-16 |
| Observations | 13,000 | 13,000 | 13,000 | 4,138 | 4,138 | 4,138 |
| Log Likelihood | 7,926.707 | 97,226.680 | 2,486.618 | 3,201.497 | 31,060.070 | 1,495.331 |
| Akaike Inf. Crit. | -15,845.410 | -194,445.400 | -4,965.236 | -6,394.995 | -62,112.150 | -2,982.661 |
| Bayesian Inf. Crit. | -15,815.520 | -194,415.500 | -4,935.345 | -6,369.683 | -62,086.840 | -2,957.349 |

**Fig. 3.**
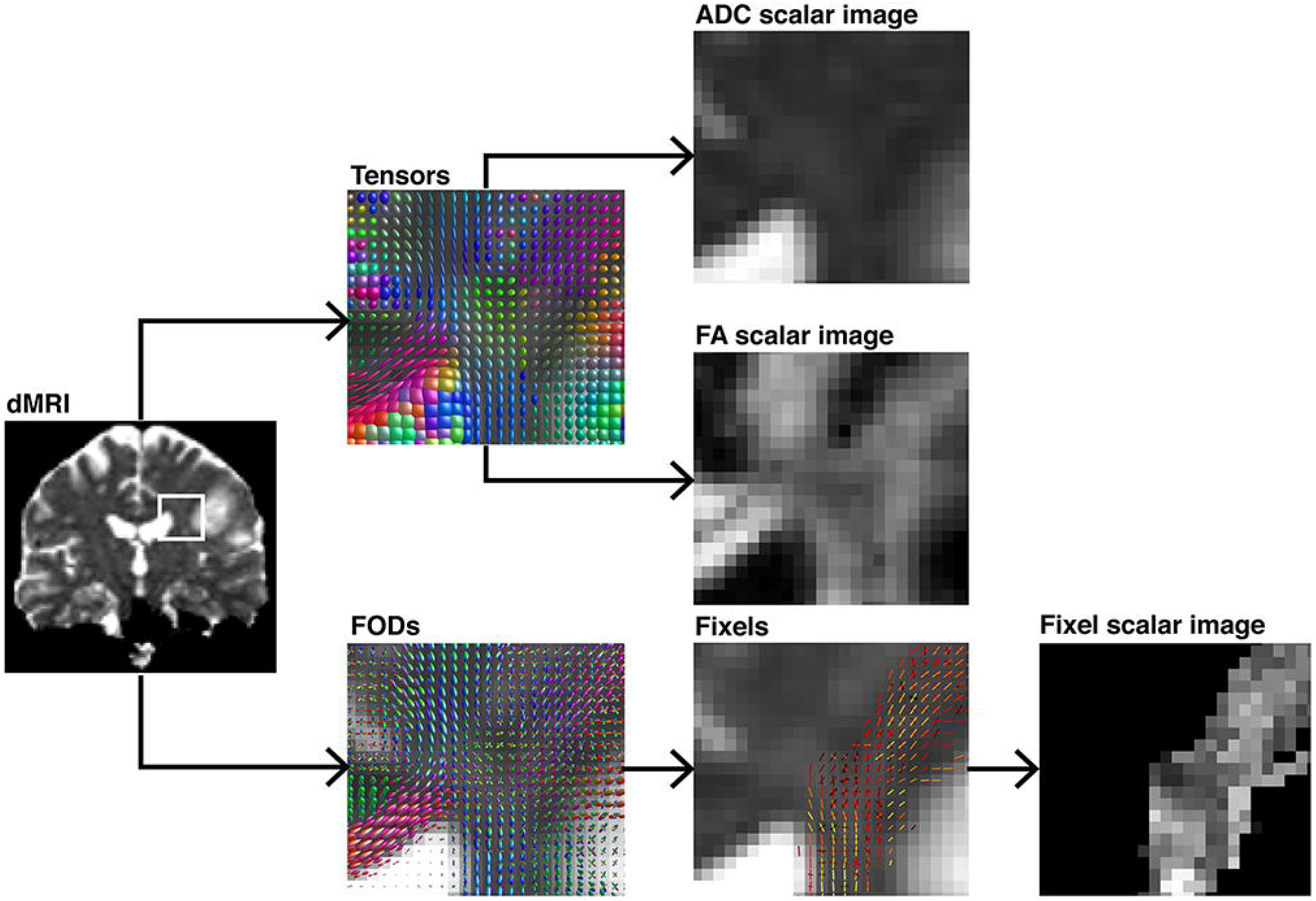
Demonstration of different voxel-level modelling methods results and their subsequently obtained scalar maps, illustrated on a coronal section. The ROI is highlighted in a pre-processed diffusion image. Either diffusion tensor-ellipsoids as estimated by diffusion tensor imaging or FOD’s estimated using CSD are shown. Further, their respective scalar maps such as ADC, FA, or fixel-based are depicted. The tensor-based scalar value does not represent any single fiber population in the voxel in comparison to the fixel-based metrics.

**Fig. 4.**
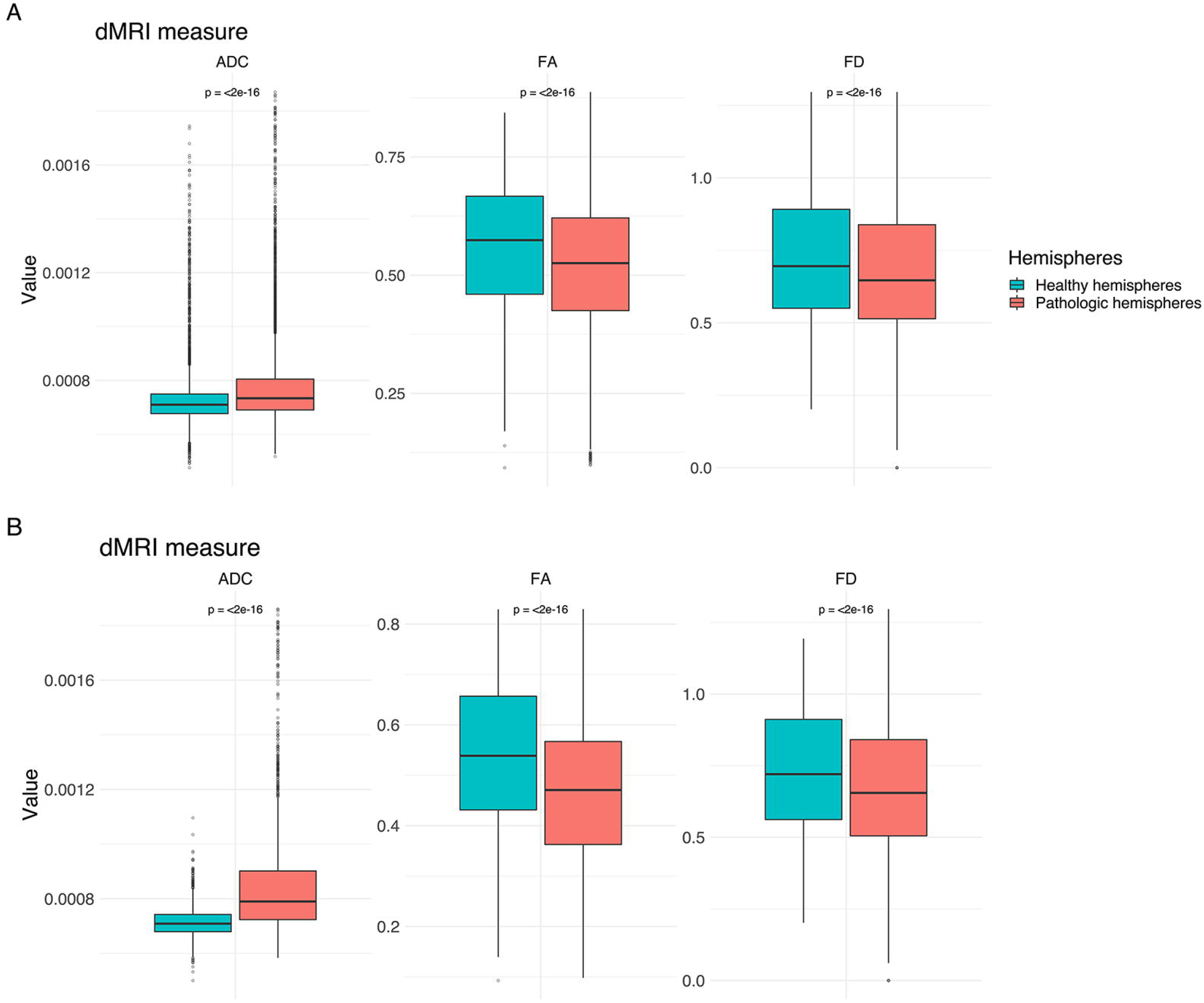
Boxplots for ADC, FA FD for both hemispheres. (A) Values for the entire CST. (B) Values for the peritumoral segments only. Outliers are marked by small circles.

**Fig. 5.**
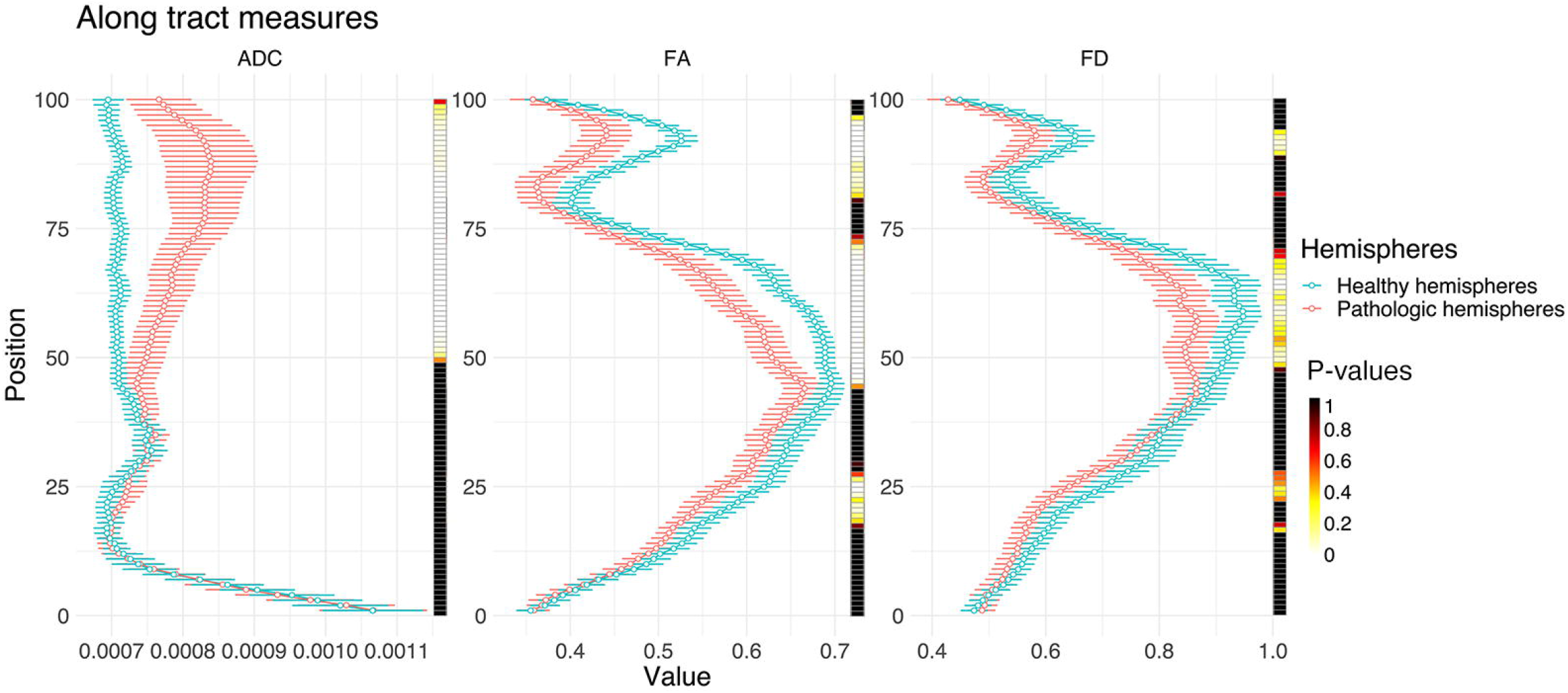
Line plots illustrating ADC, FA and FD along the entire CST of both hemispheres (0, medulla oblongata; 100, cortex). The points indicate median values with their respective 95% confidence intervals. The heat-maps demonstrate related Bonferroni-corrected p-values, derived by paired t-tests.

**Fig. 6.**
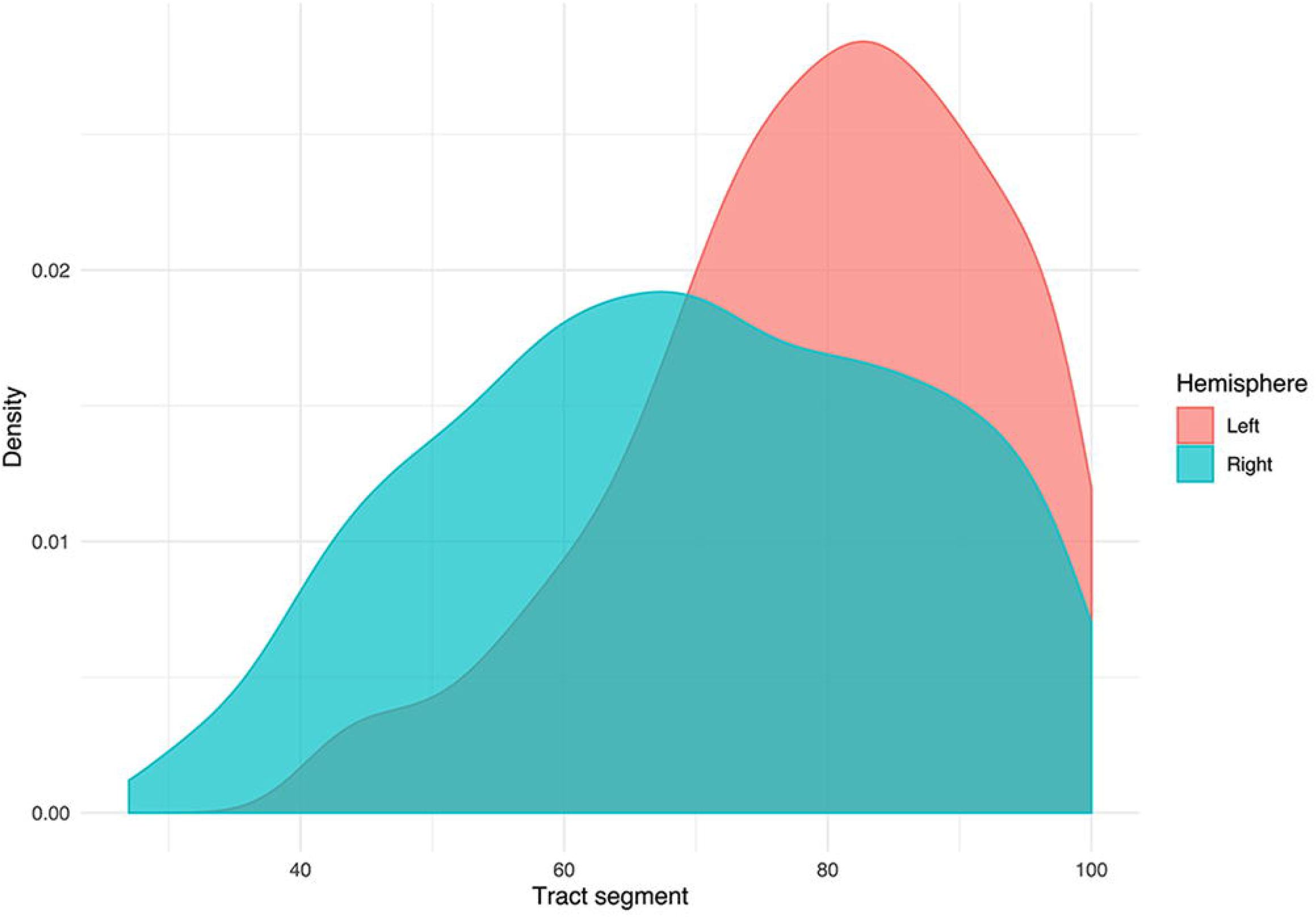
Density plot displaying the distribution of tumors grouped by hemispheric occurrence. Additionally, the plot shows that no tumors occur below segment 25.

### 3.1 Group wise analysis

The results from the mixed model analysis confirmed our hypotheses. We expected FD to improve the detection of tumor-induced changes along the tract, in combination with traditional FA or ADC measures. Furthermore, we expected stronger effects in the peritumoral segments. Our results show significant differences between healthy and pathological hemispheres for ADC, FA, and FD in the peritumoral areas (Table 2). As expected, these effects can be confirmed in the peritumoral segments in all tested values (Table 3). Fig. 4 & 5 illustrate significantly lower values in the pathological hemisphere within the entire cohort and even greater differences within the peritumoral segments for FD. Calculations for sensitivity and specificity yielded 63%, 74% and 42% sensitivity and 68%, 53% and 76% specificity for ADC, FA and FD in that order, reflecting a higher sensitivity for ADC and FA to tumor induced microstructural differences, whereas FD showed higher specificity to local WM architecture complexities or orientation dispersion.

In addition to these analyses, we calculated the mean of the entire cohort of ADC, FA and FD differences between the healthy and pathological hemispheres with respect to healthy segments only, pathological segments only and healthy-pathological WM interface (range of 3 voxels) for tumor external as well as internal segments (Fig. 7). The results indicate that ADC is stronger altered within the pathological WM area, while FA and FD show alterations along the entire CST. Furthermore, FD shows stronger differences in the healthy-pathological WM interface.

**Fig. 7.**
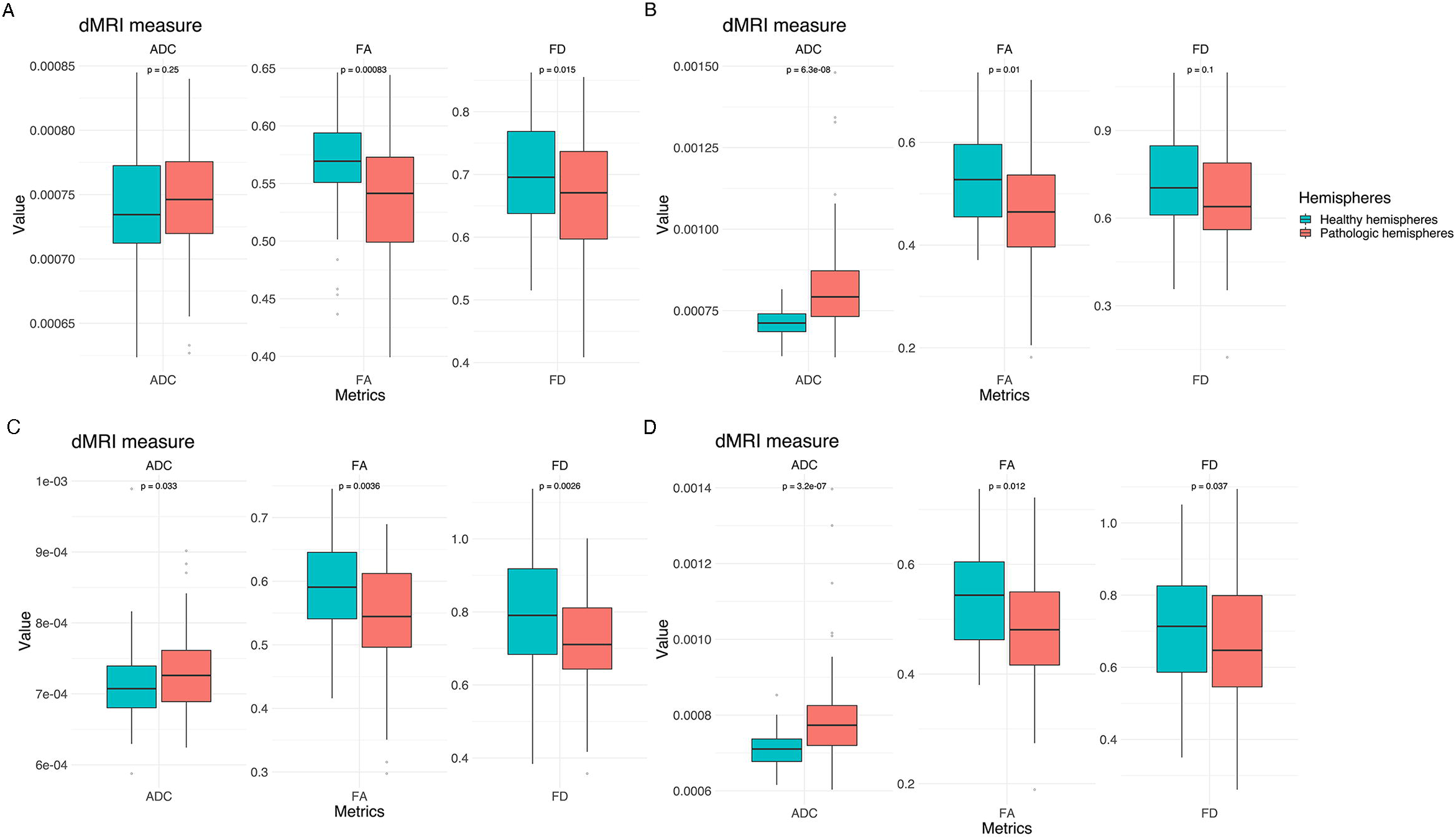
Box plots of cohort mean of ADC, FA and FD differences between the healthy and pathological hemispheres with respect to healthy segments only (A), pathological segments only (B) and tumor-healthy WM interface for tumor external (C) as well as internal segments (D).

### 3.2 Subject-specific analysis

The differences are illustrated by means of two example cases (Fig. 8 and Table 3). Four further case-specific examples are given in the supplementary materials (supplementary Fig. 1-4 & supplementary Tables 1-4). The exemplary cases were randomly selected by a script. Case A: This patient in his 80’s was brought to our emergency room with suspected stroke. A sudden weakness in the legs had occurred, causing the patient to collapse without losing consciousness. Furthermore, it was reported that the patient had been suffering from dizziness for several weeks. Conventional MRI confirmed a left parietal mass with extensive perifocal edema. The patient was diagnosed with a left postcentral WHO grade IV glioblastoma and right leg emphasized hemiparesis. The indication for resection of the mass was given.

**Table 3 (A-B).**
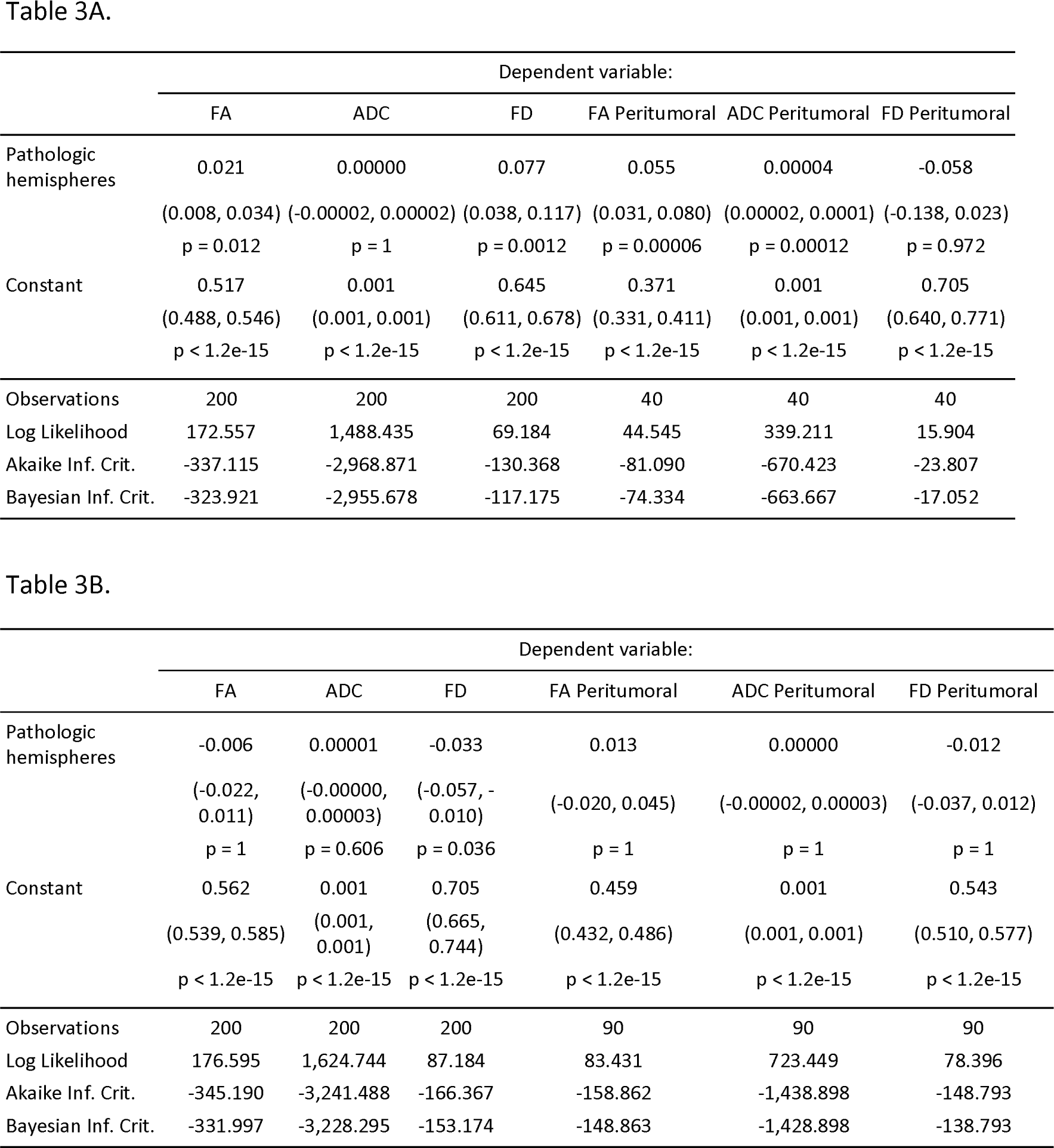
Results of linear mixed model analysis. Models 1-3 show results for the entire CST for FA, ADC, and FD, models 4-6 for the peritumoral segments respectively. The table shows regression coefficients for the fixed effect of hemisphere and the intercept with their respective standard error in brackets. Further, number of observations for each model, the log likelihood ratio, Akaike information criterion and Bayesian information criterion are stated.

**Fig. 8.**
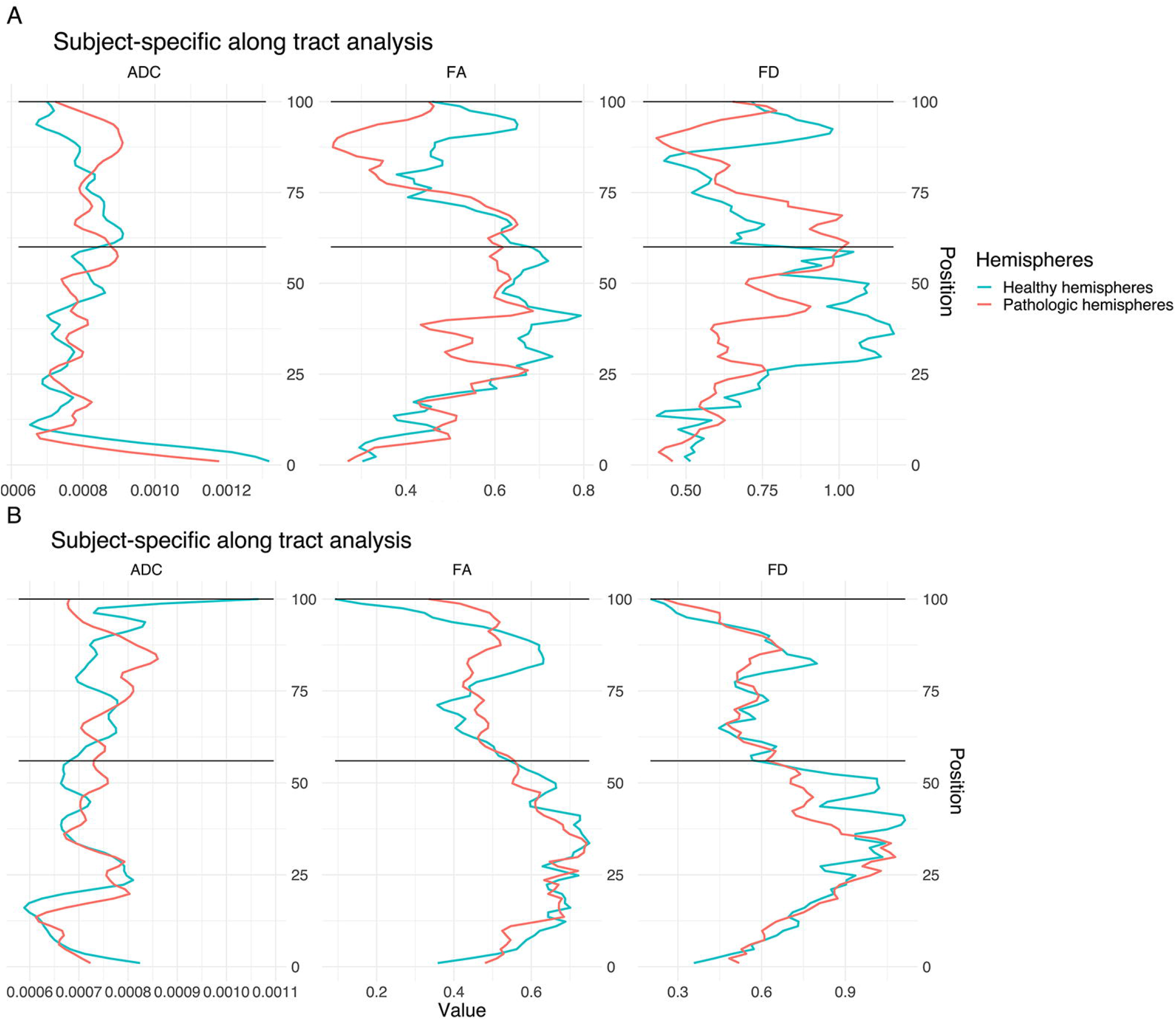
Single subject line plots depicting ADC, FA and FD along the CST of both hemispheres for case A (A) and B (B). The black lines indicate the peritumoral segments.

Case B: This patient in his 60’s presented with a several weeks history of dysesthesia in his left arm and right hand with associated arm weakness. He also felt insecure when walking and suffered from a general weakness. Conventional MRI confirmed the presence of a right frontal mass. Following this, the patient was referred to our clinic. The patient was diagnosed with a complex focal seizure with right precentral WHO grade IV glioblastoma and Todd’s paresis which included transient left hemiparesis. The indication for resection of the mass was given.

Our results show significant differences between healthy and pathological hemispheres in FD over the entire CST (p < .01 and p < .01) for both cases (Table 3). Case A shows significant differences in FA over the entire CST and in the peritumoral segments (p < .01 and p < .01). In addition, a significant difference (p < .05) can be seen in the peritumoral area as well with respect to ADC. However, case B shows no significant differences for ADC and FA, neither between the entire healthy and pathological hemispheres nor in the peritumoral segments. The values of the two hemispheres overlap here in the non-peritumoral area, similar to the group wise results described above. Case A shows less overlap for FA and FD, also in the non-peritumoral segments, while ADC shows large overlap.

## 4. Discussion

Morbidity due to brain tumor growth and their surgical treatment is often caused by impairment of relevant WM. Neuroimaging-based characterization of the healthy-pathological WM interface area is therefore crucial for neurosurgical planning. DTI based tractography has seen a widespread adoption in clinical neuroscience and practice in the recent years. Especially the combination of TMS and DTI for motor function-informed tractography has shown promising results. Yet, the interpretation of differences as measured by tensor-based scalar values is particularly challenging in regions with crossing fibers, since tensors reflect only the main diffusion direction (Jeurissen et al., 2013; D. Raffelt et al., 2012). Because the tensor representation is not able to distinguish crossing fiber populations present in the majority of the WM voxels, FA offers limited opportunities to quantitatively study WM integrity (Jeurissen et al., 2013; Van Hecke et al., 2016). Nevertheless, diffusion anisotropy can provide unique information about axonal anomalies (Mori & Tournier, 2014) as it decreases as a consequence of loss of coherence in the preferred main diffusion direction (Soares et al., 2013). In this context, studies also show that ADC is generally higher in damaged tissue due to increased free diffusion. This suggests that we can compare values of above mentioned metrics with a population average in order to determine whether they are unusually high or low, e.g. by comparing the subject-specific values of WM pathways of the healthy hemisphere with those of the pathological hemisphere or compare group-wise pathological populations with healthy ones (Mori & Tournier, 2014).

It has already been confirmed that many voxels along the CST contain considerable contributions of multiple fiber populations (Farquharson et al., 2013; Petersen et al., 2017). Nevertheless, our results indicate more significant segment-wise differences between the healthy and pathological hemispheres for FA and ADC in comparison to FD. This result was found in the group and individual tests. The investigation of other pathways may result in another order for the sensitivity and specificity of the metrics due to, for instance, different contributions of multiple fiber populations or extra axonal signal.

### 4.1 FD metrics in clinical settings

To better account for the complex microstructural organization of WM and its quantitative analysis, FD, which uses higher-order dMRI models such as FODs to analyze differences along WM pathways, allows to consider multiple fiber populations within a voxel. Multiple studies for group-wise statistical analysis of dMRI measures were published earlier (D. Raffelt et al., 2012; D. A. Raffelt et al., 2015; D. A. Raffelt et al., 2017). In contrast to these group-wise study designs, we used FD for an individual assessment of a specific tract for clinical validation. However, the presented higher sensitivity of ADC and FA indicates that these metrics are more appropriate and robust for peritumoral analysis. However, this may be due to the fact that FD has underperformed due to insufficient raw data. This finding highlights the need for better dMRI quality in clinical routine to be able to integrate advanced neuroimaging methods into clinical workflows. The discrepancy between clinical scan quality and advanced neuroimaging highlights the need to optimize raw data acquisition in order to leverage advanced neuroimaging modalities and methods into the clinical workflow (Farquharson et al., 2013; Jeurissen, Tournier, Dhollander, Connelly, & Sijbers, 2014a).

Our results demonstrate the feasibility of FD along-tract analysis as a tool to describe subject-and tract-specific tumor-induced changes. Moreover, our results demonstrate the addition of further information to that obtained only via ADC or FA. Earlier fixel studies, designed for group wise analysis of pathology-related effects, demonstrated that fixel-analyses are sensitive to WM changes in a variety of pathologies (D. A. Raffelt et al., 2015; D. A. Raffelt et al., 2017). In this study, we focused on subject-specific analyses, which showed higher sensitivity for ADC and FA, but higher specificity for FD. These findings are in line with other studies (Chamberland et al., 2019; Mormina et al., 2015). The higher specificity of FD in relation to correctly predict healthy segments is particularly relevant for presurgical analysis and intraoperative navigation in relation to risk assessment, but also for retrospective evaluation or outcome prediction models.

### 4.2 ADC, FA and FD characteristics in brain tumor patients

In both cases subject-specific differences between the healthy and pathological hemispheres can be seen in the tumorous segments. Furthermore, differences between the non-pathological and pathological area can be seen as well in non-tumorous segments. This result may indicate a global effect of gliomas on the entire CST and neural connectivity, affecting diffusion and voxel-wise white matter architecture modelling, especially in regard to FD. The results are consistent with the expected behavior of the different diffusion measures: ADC was higher in the pathological hemispheres which is attributed to the damaged tissue leading to increased diffusion. This finding might reflect the tumor-related degression of WM integrity, the edema surrounding the tumor and related increase of free-water (Mormina et al., 2015). FA and FD showed lower values in the pathological hemispheres compared to the corresponding segments in the healthy hemispheres. This result is consistent with the effect of the glioma-related loss of coherence in the preferred main diffusion directions (FA) and reduced fiber density (FD). This might be explained by the tumor infiltration or edema affecting the CST (Mormina et al., 2015). The ADC and FD values show a higher overlap of the healthy and pathological hemispheres in the non-peritumoral area.

### 4.3 Limitations

Tractography suffers from a range of limitations that make its routine use problematic (Schilling et al., 2019). It is well known that tractograms contain false positive (Maier-Hein et al., 2017) and false negative (Aydogan et al., 2018) streamlines. In addition, tractography cannot distinguish between afferent and efferent connections, and streamlines may terminate improperly (Tournier, 2019). The dMRI data used for this study consists of a typical clinical single-shell acquisition, and is thus suboptimal for fiber density measurement due to incomplete attenuation of apparent extra-axonal signal (D. Raffelt et al., 2012). In this study we focused on the CST. Further studies could integrate a variety of fiber bundles to investigate the need for FD in along-tract statistical analysis.

## 5. Conclusions

Our results show that the direct comparison between healthy and pathological hemispheres is sensitive to glioma-induced changes in structural integrity of the CST measured by different dMRI derived metrics. In contrast to our hypothesis, according to our data and analysis, FD did not outperform FA or ADC and all three metrics showed similar results for indicating tumor-induced changes of the CST. This finding highlights the need for better scans in clinical routine if one wants to introduce advanced neuroimaging modalities into clinical workflows.

## Data Availability

https://doi.org/10.5281/zenodo.3744339

## Acknowledgements

This work was supported by the DFG (EXC 2025). The views expressed are those of the author(s) and not necessarily those of the DFG. We thank Heike Schneider for the numerous TMS-mappings.

## ^1^ Abbreviations

ADC: Apparent diffusion coefficient
CSD: Constrained spherical deconvolution
CST: Corticospinal tract
dMRI: Diffusion magnetic resonance imaging
DTI: Diffusion tensor imaging
FA: Fractional anisotropy
FD: Fiber density
FDI: first dorsal interosseous
FOD: Fiber orientation distribution
GM: Grey matter
MEP: motor evoked potentials
nTMS: Navigated transcranial magnetic stimulation
RMT: Resting motor threshold
WM: White matter

