## Supplementary figures and images for "Detecting corticospinal tract impairment in tumor patients with fiber density and tensor-based metrics"

### Supplementary Fig. 1

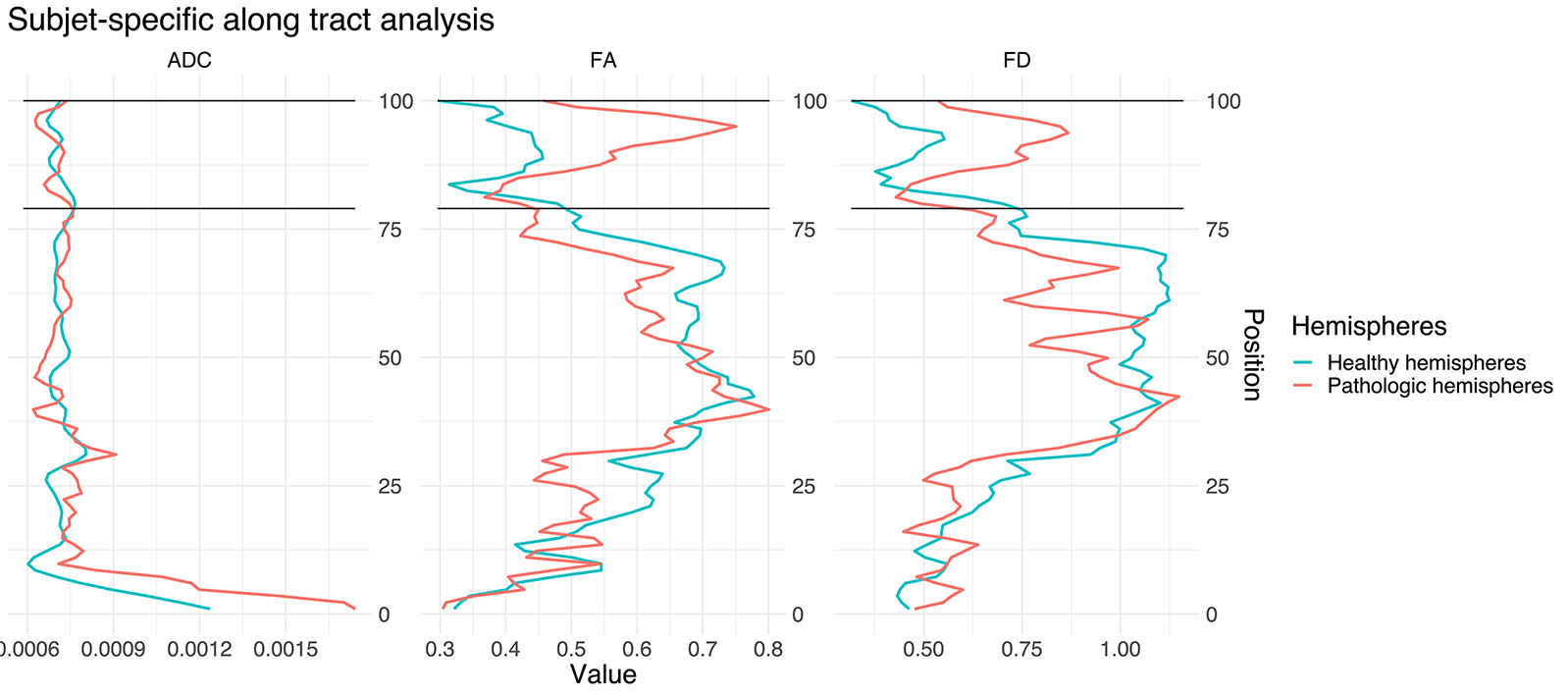

### Supplementary Fig. 2

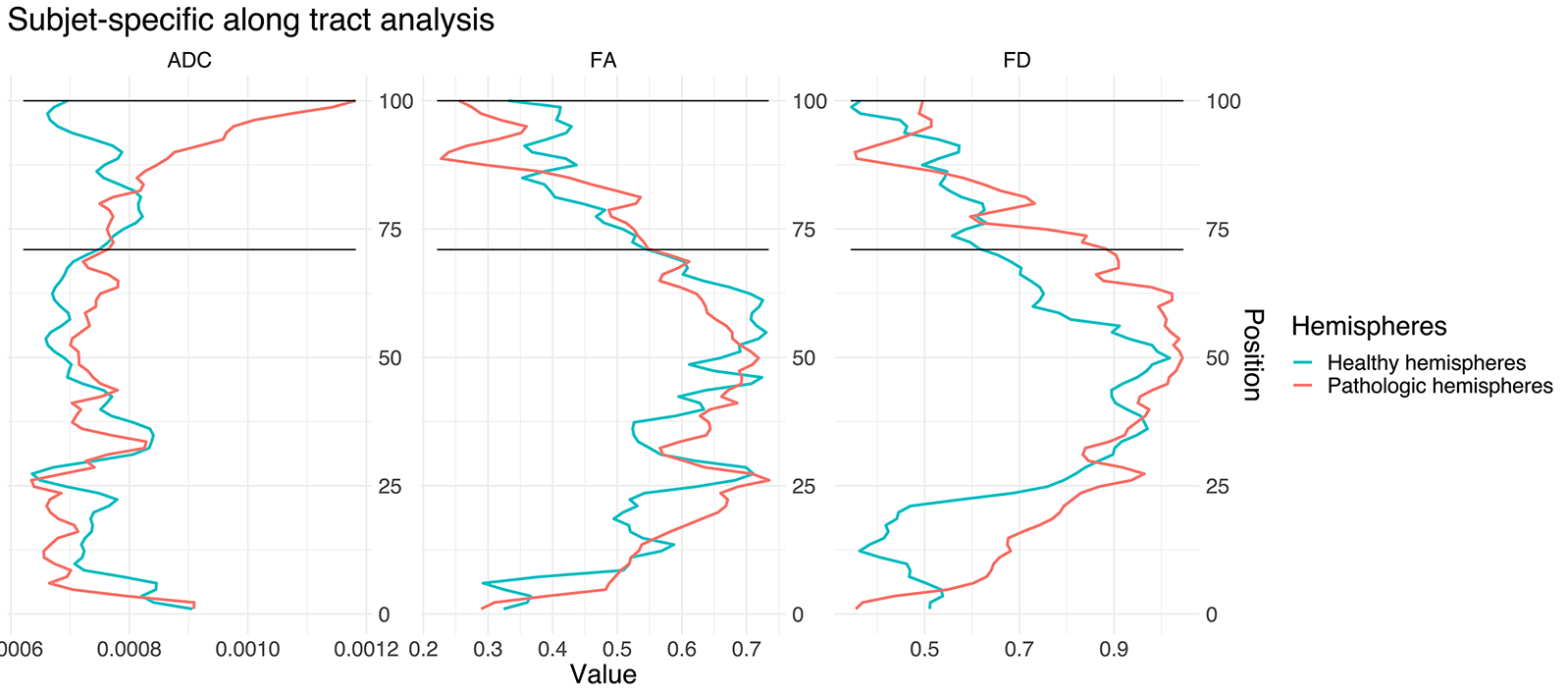

### Supplementary Fig. 3

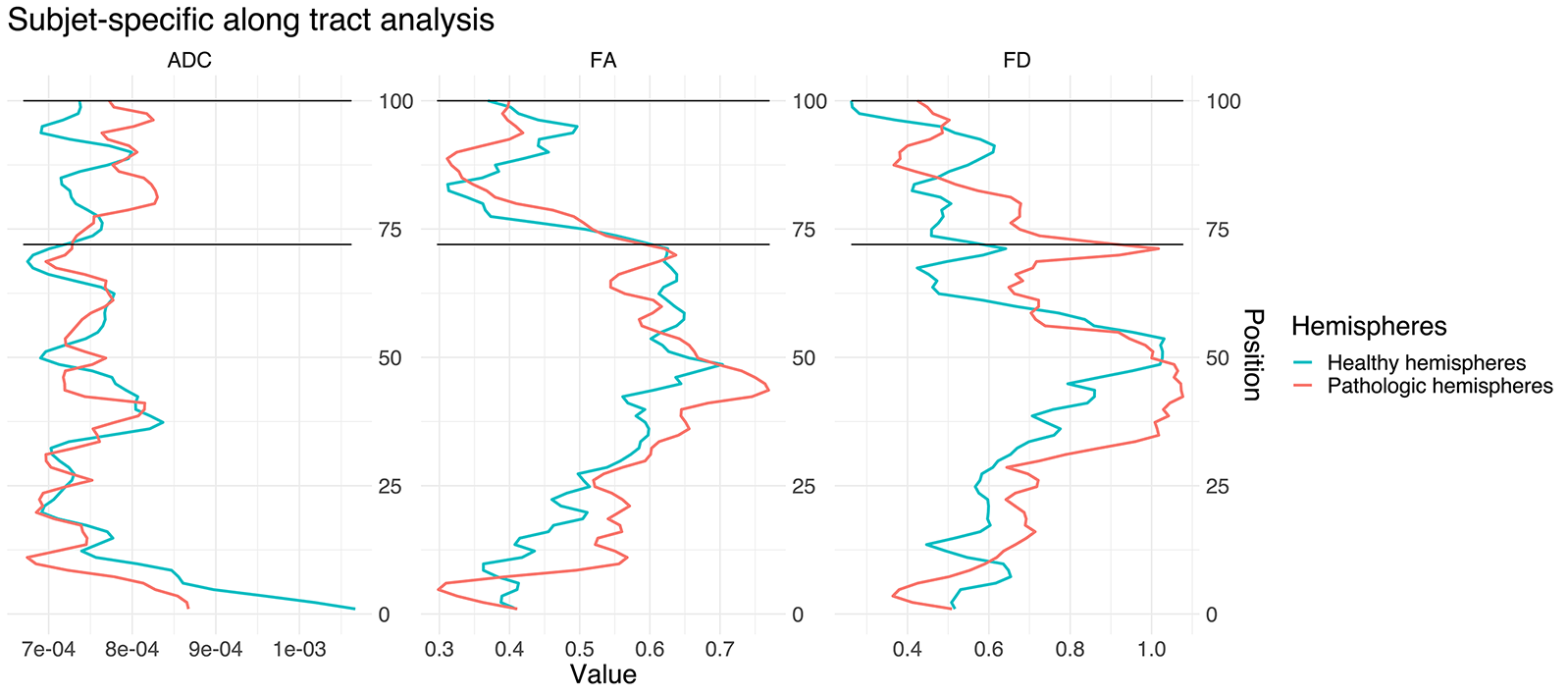

### Supplementary Fig. 4

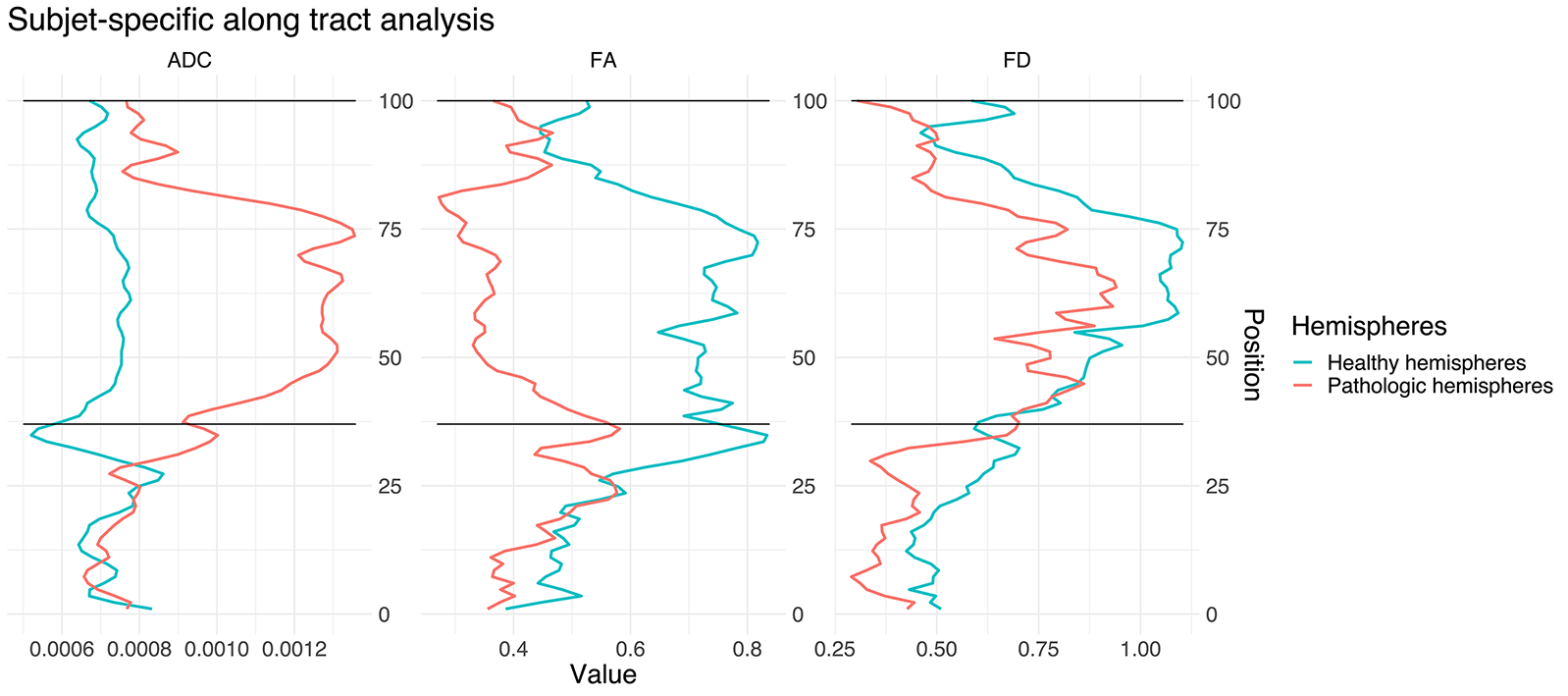
