## Supplementary Table 1-4 for "Detecting corticospinal tract impairment in tumor patients with fiber density and tensor-based metrics"

**Supplementary Table 1:** Results of linear mixed model analysis.

|  | *Dependent variable:* | | | | | |
| --- | --- | --- | --- | --- | --- | --- |
|  | FA | ADC | FD | FA Peritumoral | ADC Peritumoral | FD Peritumoral |
| Pathologic  Hemispheres | -0.011 | 0.00004 | -0.029 | 0.128 | -0.00002 | 0.169 |
|  | (-0.032, 0.010) | (0.00002, 0.0001) | (-0.065, 0.006) | (0.078, 0.178) | (-0.00003, -0.00000) | (0.093, 0.244) |
|  | p = 1 | p = 0.006 | p = 0.618 | p = 5.3e-5 | p = 0.043 | p = 0.00002 |
| Constant | 0.567 | 0.001 | 0.765 | 0.409 | 0.001 | 0.477 |
|  | (0.542, 0.591) | (0.001, 0.001) | (0.719, 0.811) | (0.370, 0.448) | (0.001, 0.001) | (0.424, 0.531) |
|  | p = 2e-16 | p = 2e-16 | p = 2e-16 | p = 2e-16 | p = 2e-16 | p = 2e-16 |
| Observations | 200 | 200 | 200 | 44 | 44 | 44 |
| Log Likelihood | 151.251 | 1,482.318 | 35.856 | 37.594 | 368.535 | 23.691 |
| Akaike Inf. Crit. | -294.501 | -2,956.635 | -63.713 | -67.188 | -729.070 | -39.383 |
| Bayesian Inf. Crit. | -281.308 | -2,943.442 | -50.519 | -60.051 | -721.934 | -32.246 |

**Supplementary Table 2:** Results of linear mixed model analysis.

|  |  | *Dependent variable:* | | | | | |
| --- | --- | --- | --- | --- | --- | --- | --- |
|  |  | FA | ADC | FD | FA Peritumoral | ADC Peritumoral | FD Peritumoral |
|  | Pathologic  Hemispheres | 0.009 | 0.00003 | 0.104 | -0.026 | 0.0001 | 0.053 |
|  |  | (-0.007, 0.025) | (0.00000, 0.0001) | (0.078, 0.130) | (-0.057, 0.005) | (0.0001, 0.0002) | (0.005, 0.100) |
|  |  | p = 0.272 | p = 0.028 | p = 0.000 | p = 0.104 | p = 0.00001 | p = 0.030 |
|  | Constant | 0.538 | 0.001 | 0.670 | 0.429 | 0.001 | 0.531 |
|  |  | (0.513, 0.564) | (0.001, 0.001) | (0.630, 0.710) | (0.397, 0.462) | (0.001, 0.001) | (0.487, 0.575) |
|  |  | p = 2e-16 | p = 2e-16 | p = 2e-16 | p = 2e-16 | p = 2e-16 | p = 2e-16 |
|  | Observations | 200 | 200 | 200 | 60 | 60 | 60 |
|  | Log Likelihood | 171.816 | 1,564.614 | 76.583 | 58.626 | 450.836 | 38.524 |
|  | Akaike Inf. Crit. | -335.632 | -3,121.228 | -145.166 | -109.252 | -893.671 | -69.049 |
|  | Bayesian Inf. Crit. | -322.438 | -3,108.035 | -131.973 | -100.875 | -885.294 | -60.671 |

**Supplementary Table 3:** Results of linear mixed model analysis.

|  | Dependent variable: | | | | | |
| --- | --- | --- | --- | --- | --- | --- |
|  | FA | ADC | FD | FA Peritumoral | ADC Peritumoral | FD Peritumoral |
| Pathologic  Hemispheres | 0.020 | -0.00000 | 0.092 | -0.012 | 0.00005 | 0.071 |
|  | (0.006, 0.034) | (-0.00001, 0.00001) | (0.062, 0.121) | (-0.035, 0.012) | (0.00003, 0.0001) | (0.010, 0.132) |
|  | p = 0.007 | p = 0.680 | p = 1.99e-08 | p = 0.328 | p = 1.19e-6 | p = 0.024 |
| Constant | 0.511 | 0.001 | 0.617 | 0.422 | 0.001 | 0.473 |
|  | (0.488, 0.533) | (0.001, 0.001) | (0.578, 0.655) | (0.394, 0.450) | (0.001, 0.001) | (0.428, 0.517) |
|  | p = 2e-16 | p = 2e-16 | p = 2e-16 | p = 2e-16 | p = 2e-16 | p = 2e-16 |
| Observations | 200 | 200 | 200 | 58 | 58 | 58 |
| Log Likelihood | 192.837 | 1,654.627 | 71.279 | 68.728 | 496.340 | 34.856 |
| Akaike Inf. Crit. | -377.673 | -3,301.253 | -134.558 | -129.457 | -984.681 | -61.711 |
| Bayesian Inf. Crit. | -364.480 | -3,288.060 | -121.365 | -121.215 | -976.439 | -53.470 |

**Supplementary Table 4:** Results of linear mixed model analysis.

|  | Dependent variable: | | | | | |
| --- | --- | --- | --- | --- | --- | --- |
|  | FA | ADC | FD | FA Peritumoral | ADC Peritumoral | FD Peritumoral |
| Pathologic  Hemispheres | -0.218 | 0.0003 | -0.150 | -0.164 | 0.0004 | -0.164 |
|  | (-0.248, -0.189) | (0.0002, 0.0003) | (-0.173, -0.128) | (-0.194, -0.134) | (0.0003, 0.0004) | (-0.194, -0.134) |
|  | p = 2e-16 | p = 2e-16 | p = 2e-16 | p = 2e-16 | p = 2e-16 | p = 1e-15 |
| Constant | 0.629 | 0.001 | 0.736 | 0.849 | 0.001 | 0.849 |
|  | (0.608, 0.650) | (0.001, 0.001) | (0.694, 0.777) | (0.803, 0.894) | (0.001, 0.001) | (0.803, 0.894) |
|  | p = 2e-16 | p = 2e-16 | p = 2e-16 | p = 2e-16 | p = 2e-16 | p = 2e-16 |
| Observations | 200 | 200 | 200 | 128 | 128 | 128 |
| Log Likelihood | 156.757 | 1,426.280 | 86.538 | 59.233 | 925.384 | 59.233 |
| Akaike Inf. Crit. | -305.514 | -2,844.560 | -165.075 | -110.467 | -1,842.768 | -110.467 |
| Bayesian Inf. Crit. | -292.320 | -2,831.367 | -151.882 | -99.059 | -1,831.360 | -99.059 |
